# A RANDOMIZED TRIAL - INTENSIVE TREATMENT BASED IN IVERMECTIN AND IOTA-CARRAGEENAN AS PRE-EXPOSURE PROPHYLAXIS FOR COVID- 19 IN HEALTHCARE AGENTS

**DOI:** 10.1101/2021.03.26.21254398

**Authors:** Rossana Elena Chahla, Luis Medina Ruiz, Eugenia Silvana Ortega, Marcelo Fabio Morales, Francisco Barreiro, Alexia George, Cesar Mancilla, Sylvia Paola D’ Amato, Guillermo Barrenechea, Daniel Gustavo Goroso, Maria de los Angeles Peral de Bruno

## Abstract

**Key Point:** *IMPORTANCE:* The emergency of COVID-19 requires the implementation of urgent strategies to prevent the spread of the disease, mainly in health personnel, who are the most exposed and has the highest risk of becoming infected with the SARS-COV-2. Drug repurposing is a pragmatic strategy, a faster and cheaper option, compared to the new drug development that has proven successful for many drugs and can be a key tool in emergency situations such as the current one that requires quick action. In addition, considering the limited access to vaccines for developing countries, preventive use of ivermectin can be a palliative that minimizes the risks of infection.

*OBJECTIVE:* To evaluate the protective effect of the combination Ivermectin / Iota-Carrageenan (IVER/IOTACRC), intensive treatment with repeated administration in oral- and nasal-spray, respectively, as a prophylaxis treatment prior to exposure to SARS-CoV-2, in health personnel at Public Healthcare Centers.

*PARTICIPANTS, DESIGN AND SETTING:* Randomized controlled 1-1 clinical trial in Personal Health, n = 234. The subjects were divided into experimental (EG: n=117; 39.6 ± 9.4 years old, 65F) and control groups (CG: n=117; 38.4 ± 7.4 years old, 61F). The EG received Ivermectin orally 2 tablets of 6 mg = 12 mg every 7 days, and Iota-Carrageenan 6 sprays per day for 4 weeks. All participants were evaluated by physical examination COVID-19 diagnosed with negative RT-PCR at the beginning, final, and follow-up of the protocol. Differences between the variables were determined using the Chi-square test. The proportion test almost contagious subject and the contagion risk (Odds Ratio) were calculated using software STATA. The level of statistical significance was reached when p-Value < 0.05.

*RESULT:* The number of subjects who were diagnosed with COVID-19 in EG was lower, only 4 of 117 (3.4%) than subjects in CG: 25 of 117 (21.4%) (*P-Value* = 1.10^−5^). Nineteen patients had mild symptoms, 4 were in EG whereas, 15 were in CG (*p*-Value = 0.001). Seven subjects were moderate, and 3 with severe diagnostics, all them in CG. The probability (Odds Ratio) of becoming ill with COVID-19 was significantly lower in EG with values of 0.13, 95% 0.03 to 0.40; *p*-Value = 1.10^−4^, this value (<1) indicates a protective effect of the IVER/IOTACRC in the EG. Logistic regression test demonstrated that treatment was effective to prevent COVID-19 (Odds Ratio 0.11, 95% 0.03 to 0.33; *p*-Value = 1.10^−4^). We also found that when increase the age, decrease contagious risk (Odds Ratio 0, 93, 95% 0.88 to 0.98, *p*-Value= 0, 02). On the other hand, the probability of contracting COVID-19 was dependent on the patient’s preexisting comorbidity (Odds Ratio 5.58, 95% 2.20 to 14.16, *p*-Value = 1.10^−5^). The other variables sex and designation were independent.

*CONCLUSION:* The intensive preventive treatment (short-term) with IVER/IOTACRC was able to reduce the number of health workers infected with COVID-19. This treatment had also effect in preventing the severity of the disease, since all patients treated were mild. We propose a new therapeutic alternative for prevention and short-term intervention scheme (intensive) that is of benefit of the health worker in this pandemic accelerated time. This intervention did not produce lack of adherence to treatment or adverse effects.

*Trial Registration:* ClinicalTrials.gov Identifier: NCT04701710

## Background

At the end of December 2019, the incidence of atypical pneumonia of unknown cause was reported in the Chinese city of Wuhan^1^. Since then, the cases have spread on a global scale generating the new COVID-19 pandemic, which represents the largest global public health crisis of this generation^2^. Genetic studies identified a new coronavirus, which was named SARS-CoV-2 due to its structural similarity with others SARS-related coronaviruses^3^.

Considering that there are no specific therapies approved by the United States Food and Drug Administration (FDA) for severe acute respiratory syndrome (SARS-CoV-2)^4^, the repositioning of different drugs with established safety profiles on the market is being studied in clinical trials and compassionate use protocols based on *in vitro* activity (against SARS-CoV-2 or related viruses) and / or on the limited clinical experience available. Drug repurposing is a pragmatic strategy, a faster and cheaper option, compared to the new drugs development that has proven successful for many drugs and can be a key tool in emergency situations such as the current one that requires quick action^5–7^. In addition, considering the limited access to vaccines for developing countries, preventive use of ivermectin can be a palliative that minimizes the risks of infection in the population.

Ivermectin is a broad spectrum anti parasitic agent approved by the FDA that in last few years has shown to have *in vitro* antiviral activity against a wide range of viruses ^4,8–11^. Caly et al. (2020) suggested that ivermectin’s nuclear transport inhibitory activity may be effective against SARS-CoV-2^12^. Different studies indicate that ivermectin would have two mechanisms of action on the COVID 19 virus: extra and intracellular. The first is through interaction with ionophores cavities or channels present in the cell membrane that electrically trap the corona of the virus capsid and prevent access to the cell^13^. The second is carried out by destabilization of the importin heterodimer complex (IMP α / β1)^13^. When destabilized, the entry to the nucleus of the virus proteins is blocked, preventing viral replication. This fact will probably result in a reduction of the antiviral responses inhibition, leading to a normal and more efficient antiviral response.

In line with these studies, numerous clinical trials are evaluating the potential of ivermectin against COVID-19 with results that are not conclusive yet regarding its efficacy and safety. At the end of March 2021, there were about 60 studies registered in https://www.clinicaltrials.gov and 43 studies listened https://www.who.int/clinical-trials-registry-platform about the safety and effectiveness of Ivermectin in COVID-19 patients, for treatment and prophylaxis^14^. A preliminary meta-analysis realized with 18 randomized Clinical Trials in 2282 patients, showed a faster time to clinical recovery and signs of viral clearance in patients who took ivermectin, comparating with control group^15^.

Carrageenans, are polysaccharides produced by algae of various families of the Rhodophyceae (red algae), its use as a food thickener additive is approved by the FDA. Its antiviral activity has been attributed to its ability to interfere with the binding of virions to host cell. Carrageenans are *in vitro* inhibitors of several viruses, including herpes simplex virus, Japanese encephalitis virus, human papilloma virus, varicella zoster virus, human rhinoviruses, and others^16^.

In this context, Health personnel are at high risk of developing the disease. Their contact with infected patients puts them at greater risk from high viral loads, resulting in more serious and prolonged illness^17–20^. Treatment with oral ivermectin, associated with iota-Carrageenan (antiviral association) applied locally in the nasal and oral cavity, would decrease the probability of the appearance or progress of clinical manifestations and the appearance of severe disease, and would decrease the viral load in the upper airway and the time of virus shedding^13^.

## Objective

The purpose of this study was to assess the effect of oral Ivermectin treatment, which has been associated with iota-carrageenan in repeated doses through the nasal and oral topical route, on the appearance and eventual progression of COVID-19 disease in a healthy population that are exposed to it and have a higher risk of contagion of SARS-COV-2 for being health personnel from community health centers, compared to standard care (usual practice).

### Primary Outcome

Reduction the infections rate for COVID-19 disease in healthcare agents.

### Secondary Outcomes

Reduction in symptoms number’s presence, and protection against the appearance of severe stages for COVID-19 disease.

## Material and Methods

### Sample Size

Sample size was determined by the test comparing two proportions^21^. It were considerate the following parameters to bilateral test: 95% confidence level, 95% statistical power, 95% proportion of infected patients in the CG, 85% proportion of infected patients in the EG. The sample size calculated, without considering losses, was 231 participants. Sample size adjusted to 20% loss ratio was 289 participants.

### Participants

The total group n = 300 to enroll included personnel who perform patient care and administrative tasks identified like: i) Healthcare: medical personnel, nurses, kinesiologists; and ii) No Healthcare: administrative and cleaning personnel. Health personnel belonging to the Tucumán State Health System (SI.PRO.SA, Tucumán, Argentina) participated in the study from October 2020 to December 2020. The recruitment procedure was managed by coordinators from each health care center who accept to participate in this trial. Enrollment was staggered until complete the sample size. The people who agreed to participate in the study gave their informed consent before starting the study (Research Ethics Committee / Health Research Directorate, file number 52/2020). The clinical trials registry number is NCT04701710. This study conforms to all CONSORT guidelines and re-ports the required information accordingly (see Supplementary Checklist).

### Inclusion criteria

Participants over 18 years of age, of both sexes, and at the start of enrollment, no subject had Covid-19 disease diagnosed by negative RT-PCR. The exclusion criteria were people under 18 years of age, pregnant or actively breastfeeding women, presenting symptoms related to COVID-19 disease, concurrent autoimmune or chronic disease, immune suppression, active infectious diseases, a history of previous SARS-CoV-2 infection confirmed by RT-PCR, medical history, and a clinical questioning.

### Design

Randomized controlled clinical trial (1:1). Once the sample was consolidated, each patient was assigned an ID corresponding to a number from 1 to 234. The selection to each group was performed through a random number generation process by an Excel spreadsheet. Then, 117 of them were randomly selected to generate the CG and EG. Figure 1 shown the consort flow diagram.

**Figure 1.**
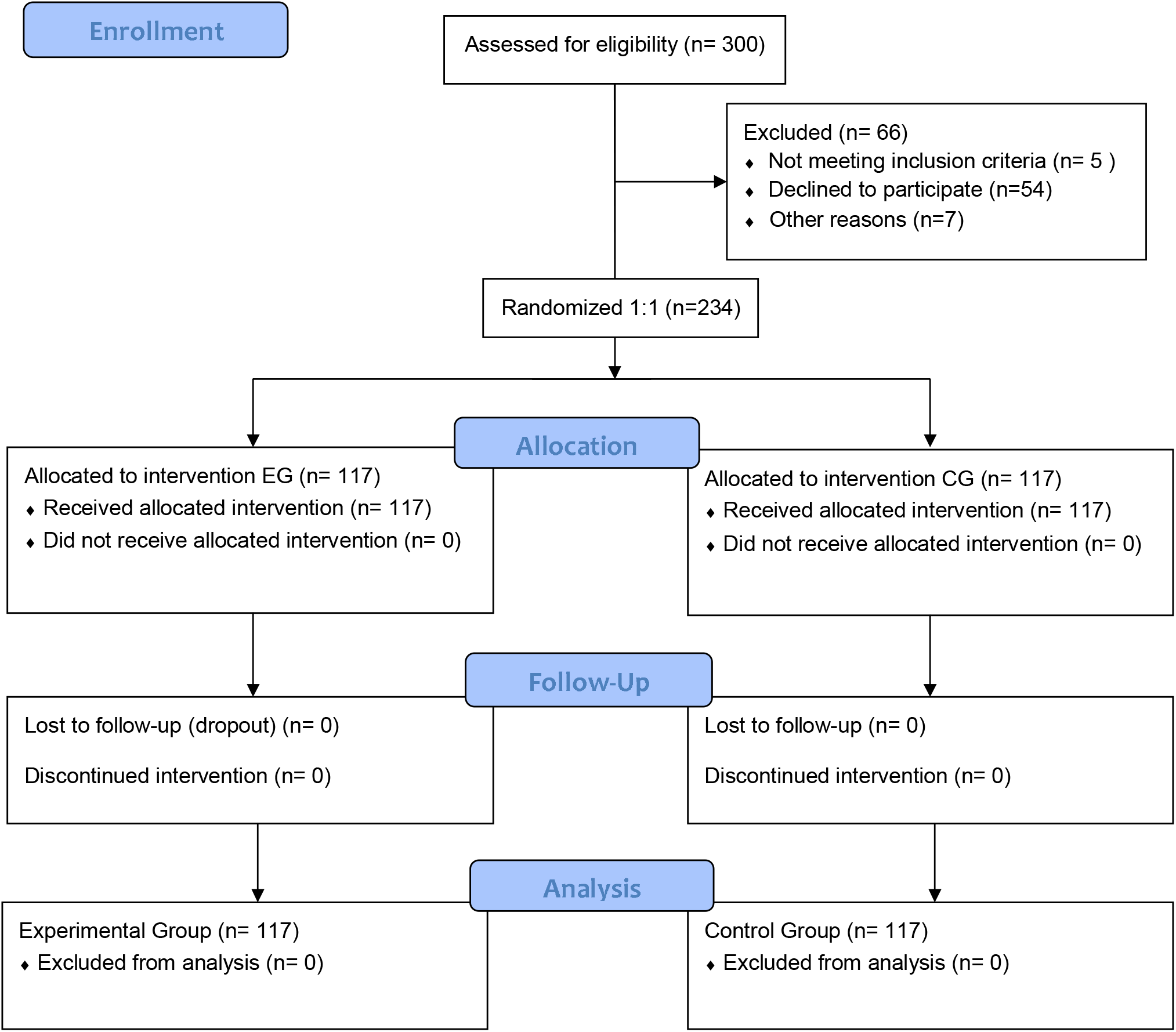
CONSORT Flow Diagram.

### Intervention Protocol

The individuals of the EG received active treatment with <u>IVER/IOTACRC</u>. Ivermectin was administered orally in 2 tablets of 6 mg = 12 mg every 7 days and Iota-Carrageenan 6 sprays per day. The entire treatment lasted 4 weeks. The CG did not receive any prophylactic treatment. Both groups used standard biosecurity care and personal protective equipment (PPE).

A post-control follow-up was carried out at 14 days (remote clinical telemedicine follow-up) at the end of which an RT-PCR test was performed. EG and CG patients were evaluated every 7 days in 4 visits from the beginning of the study. Enrolled subjects completed symptom questionnaires (including reporting any adverse effects of treatment), physical examinations, and COVID-19 nasopharyngeal secretion tests (RT-PCR) at each time. Also in the visit, in person, was supplied the corresponding dose for the week. Cases will be classified according to the WHO definitions of COVID-19 cases^22^.

### Security definitions

Adverse Event (AE) was defined as any medical event, signs, symptoms, or disease temporarily associated with the use of the medication, which could occur in the subjects enrolled in the study^23^.

### Adherence to treatment

The World Health Organization (WHO) defines adherence to treatment as compliance with it; that is, taking the medication according to the dosage of the prescribed schedule; and persistence, taking the medication over time^24^. We quantify adherence to treatment through weekly controls that include drug administration and a clinical questioning which includes the report of adverse events. Adhesion tests like Hermes, Morisky and Green have not been used, since they have been designed for treatment of chronic diseases with daily drug intake^25^. Coordinators in charge of each health care center were responsible for the recruitment and accompaniment during the trial.

### Statistics

Categorical variables were analyzed with frequencies and percentages, and continuous variables with mean and standard deviation (SD). Pearson’s Chi-square and proportions test, as appropriate, were used to analyze the statistical differences between the qualitative variables of each group. To know the contagion risk, the Odds Ratio (OR) was calculated. A Logistic Regression analysis was carried out to know the dependence between the study variables. A value of p < 0.05 was considered significant. Calculations were performed using STATA 11.2.

## Results

### Demographic profile

In total, 234 individuals from the health personnel were recruited for this study; 117 received treatment with IVER/IOTACRC and 117 within the control group who used biosecurity measures. All the participants completed the study. 57.26% of the participants enrolled in total group were women. The median age in total group was 38 years (min: 22; max: 69). 77.4% of the study participants were healthcare personnel. Table 1 shows the demographic profile and descriptions of comorbidities for the experimental and control group.

**Table 1.**
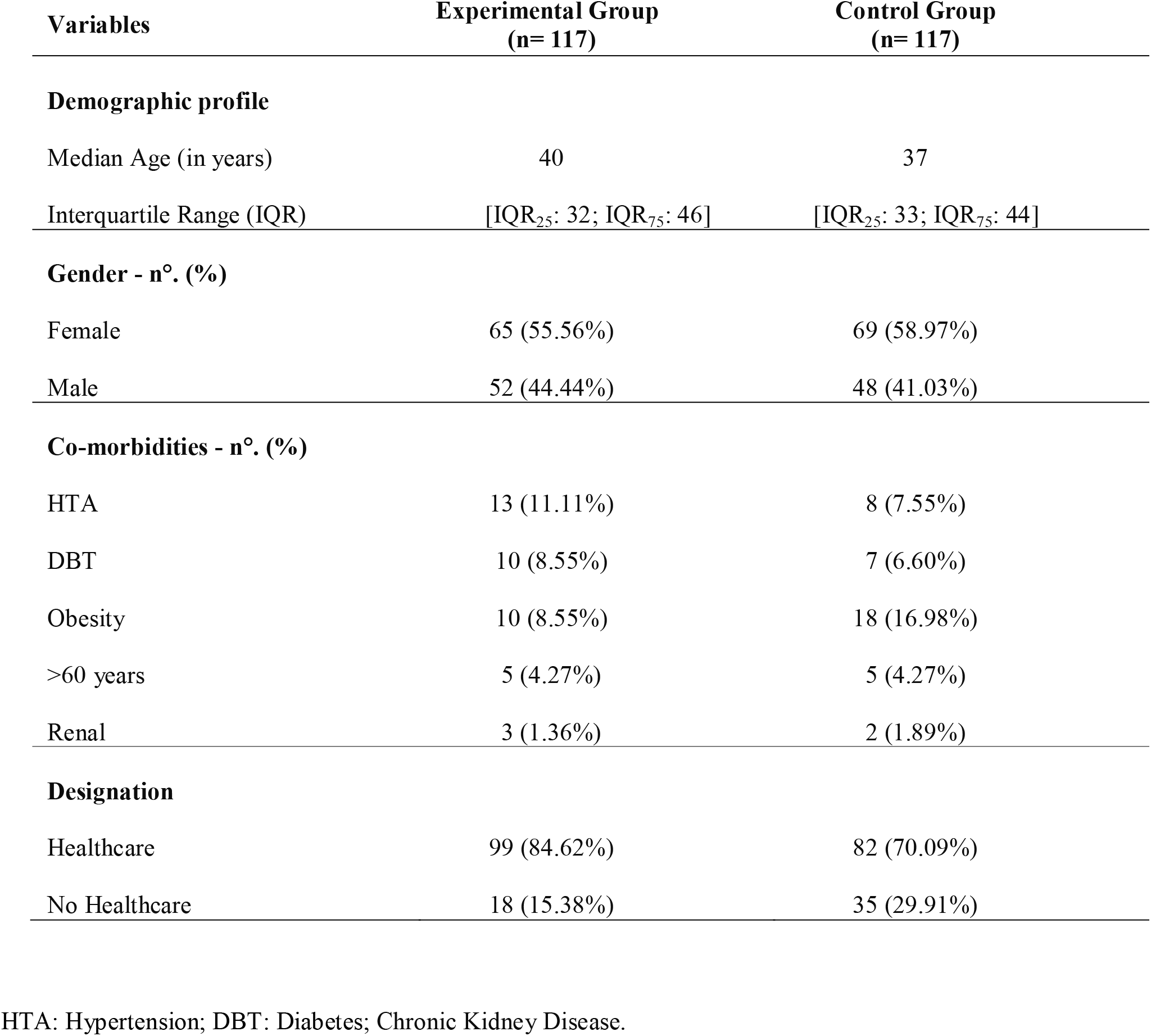
Demographic Profile

| Variables | Experimental Group<br>(n= 117) | Control Group<br>(n= 117) |
| --- | --- | --- |
| **Demographic profile** |  |  |
| Median Age (in years) | 40 | 37 |
| Interquartile Range (IQR) | [IQR<sub>25</sub>: 32; IQR<sub>75</sub>: 46] | [IQR<sub>25</sub>: 33; IQR<sub>75</sub>: 44] |
| **Gender - n°. (%)** |  |  |
| Female | 65 (55.56%) | 69 (58.97%) |
| Male | 52 (44.44%) | 48 (41.03%) |
| **Co-morbidities - n°. (%)** |  |  |
| HTA | 13 (11.11%) | 8 (7.55%) |
| DBT | 10 (8.55%) | 7 (6.60%) |
| Obesity | 10 (8.55%) | 18 (16.98%) |
| >60 years | 5 (4.27%) | 5 (4.27%) |
| Renal | 3 (1.36%) | 2 (1.89%) |
| **Designation** |  |  |
| Healthcare | 99 (84.62%) | 82 (70.09%) |
| No Healthcare | 18 (15.38%) | 35 (29.91%) |

HTA: Hypertension; DBT: Diabetes; Chronic Kidney Disease.

Table 1 shows that the demographic profile and the reported comorbidities distribution of the recruited population is homogeneous, *p*-Value> 0.05, in all the fields initially analyzed. Only, it was observed that the obese population is greater in the CG than in the EG, a relationship 18 vs. 10, respectively, with *p*-Value = 0.06 at the borderline. Similarly, the distribution of health agents in relation to their function was different in each group, after randomization was performed (*p*-Value <0.05). It should be noted that initially, no subjects had compatible COVID-19 signs, and all were diagnosed with negative RT-PCR.

### Clinical report and COVID-19 case in EG vs. CG

Table 2 shows the clinical report of the health agents after being recruited in the research. All health professionals and non-professionals were exposed to contracting COVID-19 for work reasons typical of the service.

**Table 2.**
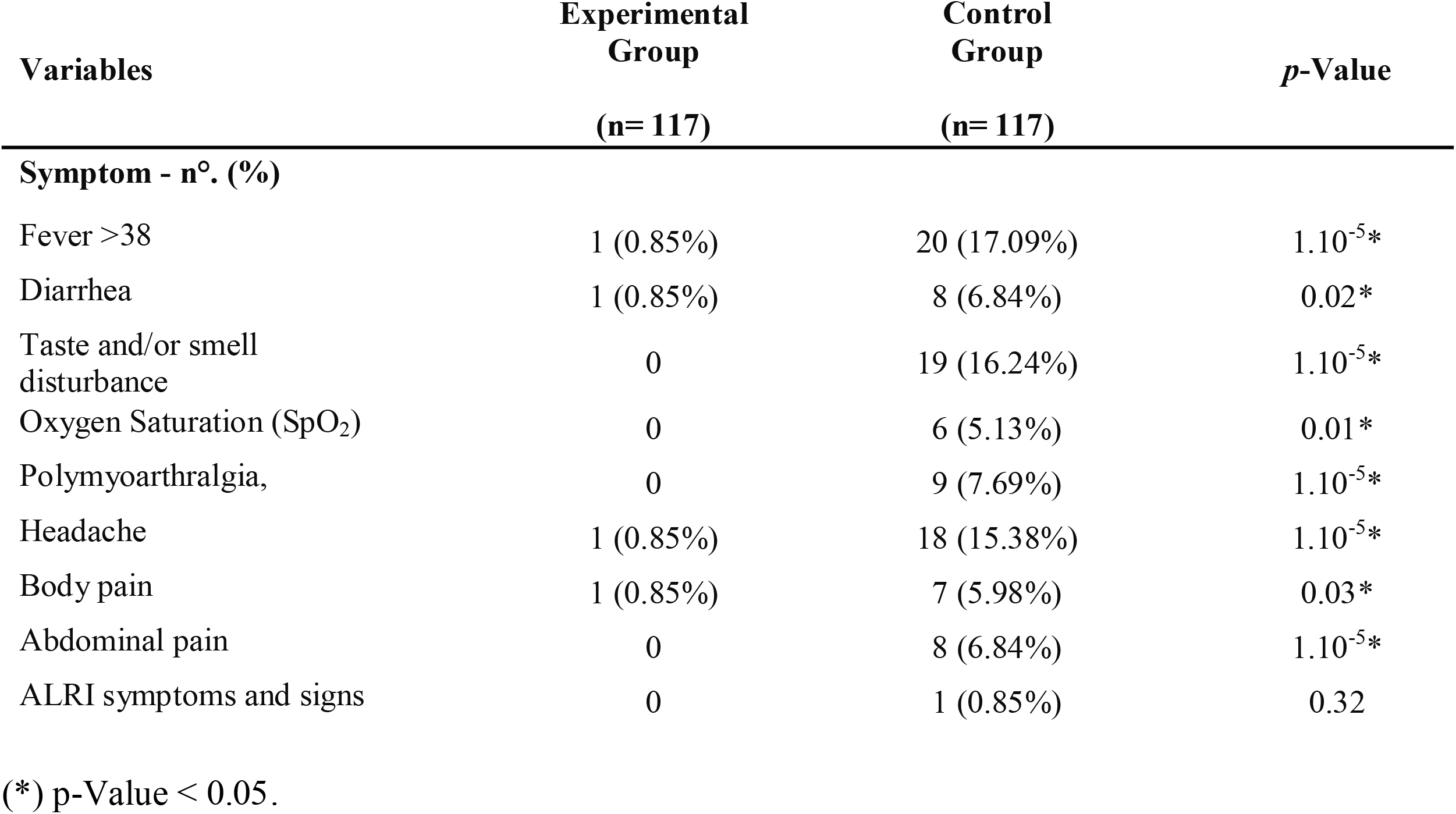
Clinical Profile

It is important to note in Table 2 that most of the symptoms, all related to COVID-19, were reported in the CG (p-Value <0.05). The most frequent symptoms were fever (21), taste and / or smell disturbance (19), and headache (19). With intermediate frequency of symptoms, cases with polymyoarthralgia (9), diarrhea (9), abdominal pain (8), and low oxygen saturation (SpO_2_) (6) were reported. Symptoms related to ALRI symptoms and signs (1) were reported with lower frequencies. Table 2 shows the significant differences (*p*-Value < 0.05) between EG vs CG in relation to each of the reported symptoms. CG had a prevalence of all the most frequent symptoms in people who acquired COVID-19.

Figure 2A shown that the number of subjects who were diagnosed with COVID-19 in EG was lower, only 4 of 117 (3.4%), than subjects in CG: 25 of 117 (21.4%) (*p*-Value = 1.10^−4^). Patients diagnosed with COVID-19 were classified as mild, moderate and severe, according to the gravity cases. Figure 2B shows the distribution of cases in each group and their respective classification.

**Figure 2.**
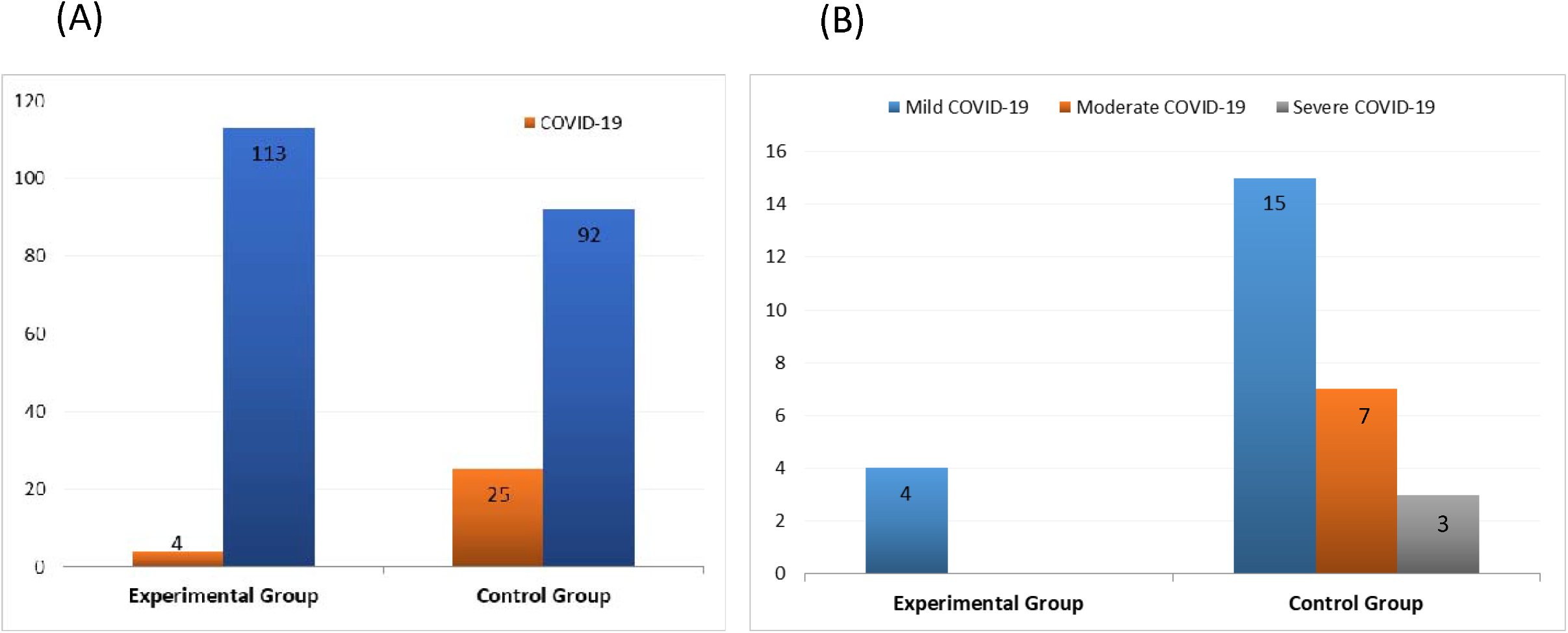
COVID-19 case in EG vs CG. A) Number of COVID-19 and healthy cases in Experimental and Control Group (n=234). B) Clinical state of the COVID-19 cases in Experimental and Control Group (n=234).

Nineteen patients had mild classification for COVID-19, n= 4 in EG, and n= 15 in CG (*p*-Value = 0.001). Seven subjects were moderate, and 3 with severe diagnostics, all them were in CG. In addition, it was found that in the EG people who contracted COVID-19 only 1/4 had any symptoms, while the CG 24/25 (*p*-Value = 1.10^−5^).

### Odds Ratio and variables influence on intervention

The probability (Odds Ratio) of becoming ill with COVID-19 was significantly lower in EG with values of 0.13, 95% 0.03 to 0.40; *p*-Value = 1.10^−4^, than in GC with values of 7.67, 95% 2.57 to 22.85; *p-*Value = 1.10^−4^. The value <1 indicates a protective effect of the IVER / IOTACRC for EG. Consequently, people with treatment decrease their chance of contracting COVID-19 by 87%.

Logistic regression test was also performed in order to determinate the influence of different variables on the clinical trials. In this model dichotomous dependent variable was used as having or not suffering from COVID-19 in relation to the five variables: IVER/IOTACRC intervention, comorbidity, age, sex and designation. Figure 3 shows the influence of different variables on the probability to getting or not COVID-19. The probability (Odds Ratio) in relation at all variables was that becoming ill with COVID-19 was maintained significantly lower in people treated with IVER/IOTAC relative to non-treated people, Odds Ratio 0.11, 95% 0.03 to 0.33; *p*-Value = 1.10^−4^. We find that the mean value, including the Confidence Interval (CI), was <1. This value indicates that the protective effect of the IVER/IOTACRC in relation to the relative reduction of the risk to contracting COVID-19 were maintained even in interaction with other variables.

**Figure 3.**
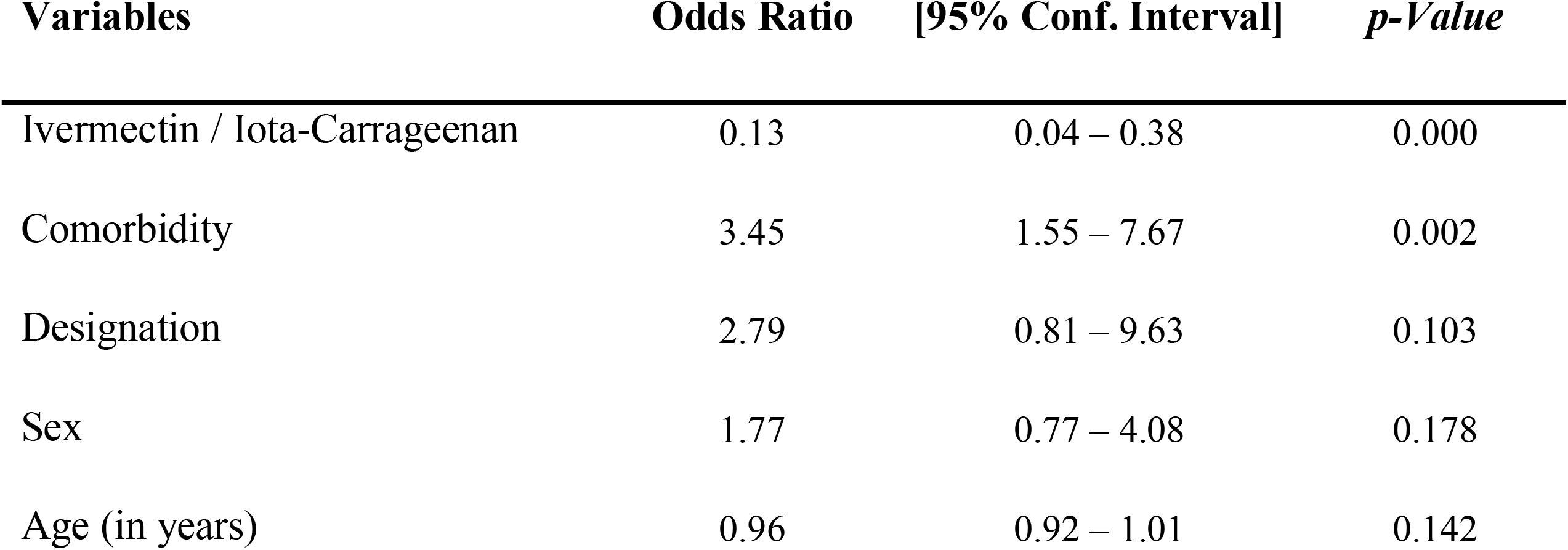
Logistic regression model in patient with COVID-19 in both groups.

**Table 3**. Influence of different variables on the probability to getting or not COVID-19.

On the other hand, the probability of contracting COVID-19 was dependent on the patient’s preexisting comorbidity. People with comorbidities had a greater chance to contracting COVID-19, Odds Ratio 5.58, 95% 2.20 to 14.16, *p*-Value = 1.10^−5^ (Odds Ratio >1).

Regarding to age, this was study as continues variable, it can be observed that as this increase, they had minor chance of getting COVID-19. This indicates that as age increases by one unit, the chance of getting or contracting COVID-19 decreases 7% the chance of getting COVID-19. This is because the Regression Coefficient (RC) has a negative sign (RC = −2.37, 95% −0.12 to −0.01, *p*-Value = 0.018). This may be due to the fact that the average age of all people enrolled in this study was 39 years, no significative differences in booth EG and CG groups (Table 1), range between 32 to 41 year was 48.3%. Odds Ratio to this variable was 0.93, 95% 0.88 to 0.98, *p-*Value = 0.02.

Getting COVID-19 was independent of sex when this variable was analyzed in both groups (CG and EG) (Table 3). When this variable was studied using a stratified model in male and female (see Table 1), we founded that the protective power of ivermectin is conserved in both sex groups (Sex F Odds Ratio 0.148, 95% 0.02 to 0.55 *p*-Value = 0.0012 Sex M Odds Ratio 0.098, 95% 0.002 to 0.796; *p-*Value = 0.010)

When the variable was studied using a stratified model in four age interquartile (see Table 1), we founded the protective power of ivermectin is conserved in the first three age interquartiles. In people older 45 years of age we found the preventive treatment wasn’t effective.

In relation to designation (Healthcare vs. no-Healthcare) and comorbidities getting COVID-19 was independent of this variable (see Table 3).

## Discussion

Health personal is one of the most exposed groups to COVID-19 contagion, because of their steady contact with infected patients. In our work we found a protective effect of the intensive IVER/IOTACRC treatment in pre-exposure prophylaxis to COVID-19 in health agents. The number of people affected by the disease was significantly higher in the CG when compared to the EG who followed the intervention. In agreement with our findings, Tarek Alacom et al. in an observational prophylactic study conducted in 118 healthcare workers, they found that significant minor contagious in subjects which received ivermectin^26^. In the aforementioned study, a lower dose of ivermectin was used unlike the treatment proposed here, held for one month and iota carrageenan was used in conjunction with ivermectin. The findings in our work, in agreement with Carvallo H et al., confirm the hypothesis that the association IVER/IOTACRC works by decreasing the possibility of infection with SARS-CoV-2 and possibly acts synergistically^27^. We interpret that a double viral barrier would be formed that would enhance its action and allow to increase the protective effect in the following way: i) The first barrier for viral protection would be at the entry of the virus into the nasal cavity where the carrageenan would behave as a mucolytic agent in the barrier of sulfacted polysaccharides with negative charge^28^; ii) The other action of ivermectin is to decrease the viral load based on its systemic cellular action^29^. It is coincident with reports of viral clearance in other clinical trials which evaluate the use of ivermectin to treat COVID-19. Ahmed S. et al. found that a 5-day course of ivermectin resulted in earlier clearance of the virus compared to placebo group ^30^.

It is understood that it is capable of preventing the entry into the cell nucleus of the viral RNA by blocking importin alpha/beta, thereby preventing replication since SARs-COVID-2 does not have the nuclear mechanisms and enzymatic actions for the transcription of new viral replicates^31^. In this direction, our work meets the work of Sharun et al (2020)^32^, who demonstrated the effect of ivermectin as a drug for inhibiting virus replication *in vitro* laboratory conditions and places the drug as a new therapeutic candidate against SARS-CoV-2 / COVID-19. There are other works, either in prevention, that found that a two-dose of ivermectin was associated with a reduction of SARS-CoV-2 infection, what makes ivermectin useful for healthcare personal preventive use^33^.

Secondary outcome found was that IVER/IOTACRC not only prevents the infections rate, but also has a protective effect on reduction in symptoms number’s presence, and protection against the appearance of severe stages for COVID-19 disease (Figure 2). As can be seen in Figure 2B, the EG only had mild cases, while the CG had mild, moderate and severe cases, the differences between both groups being significant. We observed in Table 2 that the symptoms description in the EG is significantly lower that CG. On the other hand, it’s necessary point out that the comorbidities or risk factor such as hyper-tension, DBT, obesity or over 60 years old were similar in booth group (Table 1). So, the results above mentioned, cannot attributed to presence to comorbidities in the CG. In our greatest consideration, this would be an important contribution. When the effectiveness of IVER / IOTACRC treatment was analyzed together with the other variables, we found that, even in the presence of the comorbidity variable, the protective effect of IVER / IOTACRC was maintained, with Odds Ratio <1 (Table 3). It is observed that the protective effect only has a small and no significant increased (87% to 89%) in the chance getting COVID-19 in EG.

In relation to comorbidities and their greater impact on the severity of COVID-19, other researchers have shown a positive relationship^34,35^. In relation to the sex variable, in total group, we found that was independent in relation to treatment with IVER/IOTACRC (Table 3). Stratified model by age showed that treatment was protective for people under 45 years old, independent of sex.

The proposed prophylactic treatment is also independent of the designation (healthcare and no healthcare).

During the study, there was no lack of adherence to IVER/IOTACRC treatment. We hypothesized that good adherence was due to the design of the protocol, since it provided for the follow-up of the enrolled subjects periodically. These were designed every seven days using two strategies: i) face-to-face visits, and ii) remote monitoring via telemedicine. Another fact that may have influenced good adherence is that a short-term intensive protocol was used.

### Adverse effects

Regarding adverse effects, they were not reported in any case. The explanation for this is that it could be due to the fact that IVER/IOTACRC only produces these effects when the drug acts as an anti parasitic, unlike the viricidal action proposed in this study. Another fact that reinforces the absence of adverse effects is that the doses used in this protocol are low doses, in which previously, in the literature, it has been reported that they do not produce adverse effects^37^.

### Benefits

Through this study, it was possible to show a prophylactic effect of IVER/IOTACRC against COVID-19 disease. This association of drugs was inexpensive and is also accessible in the local pharmaceutical industry (Argentina). It is more relevant considering the limitations in vaccines supplies.

### Limitations

The main limitation of this study was the number of agents to enroll. This trial does not include the report of adverse event in the long run, so will be interesting to include in future trials biochemical examination for control of potential adverse effects. Financial limitations impacted in the study design, which not involved blinded evaluation and/or placebo administration. It’s also for considering the limitations of RT-PCR test in relation to diagnosis, which in future works can be complimented with other approved qualitative tests. On this last point, economic constraints had a determining rol.

### Future work

We consider that our results, taking together with other trials, are encouraging for develop further studies. New clinical intervention studies in our region and also partners in other countries that may show the effect of the IVER/IOTACRC compound in mildstage outpatients. The design that would be proposed would be to use the same treatment time but at higher doses. Other way, more strong results could be obtained from randomized double blinded studies with long term controls to arrive to solid conclusions about safety and efficacy of IVER/IOTACRC

## Conclusion

The intensive preventive treatment (short-term) with IVER/IOTACRC was able to reduce the health workers number infected with COVID-19. This treatment had an additional effect in preventing the severity of the disease, since most of the patients who received the treatment were mild.

In the presence of the comorbidity variable, the protective effect of IVER / IOTACRC was maintained in the chance getting COVID-19 in the treatment group.

The proposed prophylactic treatment is independent of the sex variable, and designation (healthcare and no healthcare).

We propose a new therapeutic alternative for prevention and short-term intervention scheme (intensive), which is of benefit of the health worker in this pandemic accelerated time. This intervention did not produce lack of adherence to treatment or adverse effects.

## Data Availability

We put at disposal the data used for those who request them

## Authors’ contributions

ESO supervised the database. ESO and DGG contributed with the data processing and contributed to the statistical analysis. ESO, DGG and MPB were responsible for writing the manuscript. MFM, FB, AG, CM and SPB contributed to data collection. REC and LMR were the institutional managers to carry out the work. MPB supervised the project.

## Transparency Declaration

The authors not receive any monetary compensation for this work. They declare that they have no known competing financial interests or personal relationships that could have appeared to influence the work reported in this paper.

## Acknowledgements

All the authors are grateful for the collaboration of the health and administrative personnel of the “Angel C. Padilla” Clinical Hospital, “Nicolás Avellaneda” Hospital, and the Emergency Department 107, and particularly, to Dr. Rogelio Calli contribution, Director of Epidemiology of the Tucumán Province, Argentina. The authors thank Dr. Augusto Bellomio (INSIBIO CONICET-UNT) for critical reading of the manuscript.

## References

1. WHO report 2020. WHO issues consensus document on the epidemiology of SARS.

2. Wang W, Xu Y, Gao R, et al. Detection of SARS-CoV-2 in Different Types of Clinical Specimens. JAMA. March 2020. doi:10.1001/jama.2020.3786

3. Wu Z, McGoogan JM. Characteristics of and Important Lessons From the Coronavirus Disease 2019 (COVID-19) Outbreak in China. JAMA. 2020;323(13):1239. doi:10.1001/jama.2020.2648

4. González Canga A, Sahagún Prieto AM, Diez Liébana MJ, Fernández Martínez N, Sierra Vega M, García Vieitez JJ. The Pharmacokinetics and Interactions of Ivermectin in Humans—A Mini-review. AAPS J. 2008;10(1):42–46. doi:10.1208/s12248-007-9000-9

5. Chong CR, Sullivan DJ. New uses for old drugs. Nature. 2007;448(7154):645–646. doi:10.1038/448645a

6. Liu Z, Fang H, Reagan K, et al. In silico drug repositioning-what we need to know. Drug Discov Today. 2013;18(3-4):110-115. doi:10.1016/j.drudis.2012.08.005

7. Ashburn TT, Thor KB. Drug repositioning: identifying and developing new uses for existing drugs. Nat Rev Drug Discov. 2004;3(8):673–683. doi:10.1038/nrd1468

8. Götz V, Magar L, Dornfeld D, et al. Influenza A viruses escape from MxA restriction at the expense of efficient nuclear vRNP import. Sci Rep. 2016;6(1):23138. doi:10.1038/srep23138

9. Wagstaff KM, Rawlinson SM, Hearps AC, Jans DA. An AlphaScreen ® -Based Assay for High-Throughput Screening for Specific Inhibitors of Nuclear Import. J Biomol Screen. 2011;16(2):192–200. doi:10.1177/1087057110390360

10. Lundberg L, Pinkham C, Baer A, et al. Nuclear import and export inhibitors alter capsid protein distribution in mammalian cells and reduce Venezuelan Equine Encephalitis Virus replication. Antiviral Res. 2013;100(3):662–672. doi:10.1016/j.antiviral.2013.10.004

11. Tay MYF, Fraser JE, Chan WKK, et al. Nuclear localization of dengue virus (DENV) 1-4 non-structural protein 5; protection against all 4 DENV serotypes by the inhibitor Ivermectin. Antiviral Res. 2013;99(3):301–306. doi:10.1016/j.antiviral.2013.06.002

12. Caly L, Druce JD, Catton MG, Jans DA, Wagsta KM. The FDA-approved drug ivermectin inhibits the replication of SARS-CoV-2 in vitro. Antiviral Res. 2020;178(January):104787.

13. Padhy BM, Mohanty RR, Das S, Meher BR. Therapeutic potential of ivermectin as add on treatment in COVID 19: A systematic review and meta-analysis. J Pharm Pharm Sci. 2020;23:462–469. doi:10.18433/jpps31457

14. Cassará FP. Ivermectina asociada a iota-Carragenina aplicada localmente en la cavidad bucal, en la profilaxis de la enfermedad COVID-19 en el personal de salud. Estudio IVERCAR01. 2020:1–23.

15. Hill A, Abdulamir A, Ahmed S, et al. <p>Meta-analysis of randomized trials of ivermectin to treat SARS-CoV-2 infection</p>. 2021:1–37.

16. Pacheco-Quito E-M, Ruiz-Caro R, Veiga M-D. Carrageenan: Drug Delivery Systems and Other Biomedical Applications. Mar Drugs. 2020;18(11):583. doi:10.3390/md18110583

17. Andersen KG, Rambaut A, Lipkin WI, Holmes EC, Garry RF. The proximal origin of SARS-CoV-2. Nat Med. 2020;26(4):450–452. doi:10.1038/s41591-020-0820-9

18. Recalcati S. Cutaneous manifestations in COVID□19: a first perspective. J Eur Acad Dermatology Venereol. 2020;34(5). doi:10.1111/jdv.16387

19. Mehta P, McAuley DF, Brown M, Sanchez E, Tattersall RS, Manson JJ. COVID-19: consider cytokine storm syndromes and immunosuppression. Lancet. 2020;395(10229):1033–1034. doi:10.1016/S0140-6736(20)30628-0

20. Chen G, Wu D, Guo W, et al. Clinical and immunological features of severe and moderate coronavirus disease 2019. J Clin Invest. 2020;130(5):2620–2629. doi:10.1172/JCI137244

21. Glueck DH. Sample Size Calculations in Clinical Research 2nd edition by CHOW, S.-C., SHAO, J., and WANG, H. Biometrics. 2008;64(4):1307–1308. doi:10.1111/j.1541-0420.2008.01138_10.x

22. World Health Organization. Clinical Spectrum of SARS-CoV-2 Infection. https://www.covid19treatmentguidelines.nih.gov/overview/clinical-spectrum/. Published 2020.

23. WHO-adverse reaction terminology (WHO-ART). In: Dictionary of Pharmaceutical Medicine. Vienna: Springer Vienna; 2009:192–193. doi:10.1007/978-3-211-89836-9_1467

24. Osterberg L, Blaschke T. Adherence to Medication. N Engl J Med. 2005;353(5):487–497. doi:10.1056/NEJMra050100

25. Puigdemont N, Valverde I. Methods to assess medication adherence. Ars Pharm. 2018;59(3):163-172. %0Ascielo.isciii.es/pdf/ars/v59n3/2340-9894-ars-59-03-163.pdf%0A%0A.

26. Alam MT, Murshed R, Gomes PF, et al. Ivermectin as Pre-exposure Prophylaxis for COVID-19 among Healthcare Providers in a Selected Tertiary Hospital in Dhaka – An Observational Study. Eur J Med Heal Sci. 2020;2(6). doi:10.24018/ejmed.2020.2.6.599

27. Hector E C. Covid 19 and Ivermectin Prevention and Treatment Update. J Infect Dis Travel Med. 2020;4(S1). doi:10.23880/jidtm-16000s1-007

28. Héctor C, Roberto H, Psaltis A, Veronica C. Study of the Efficacy and Safety of Topical Ivermectin + Iota-Carrageenan in the Prophylaxis against COVID-19 in Health Personnel. J Biomed Res Clin Investig. 2020;2(1). doi:10.31546/2633-8653.1007

29. Krolewiecki A, Lifschitz A, Moragas M, et al. Ivermectin as Pre-exposure Prophylaxis for COVID-19 among Healthcare Providers in a Selected Tertiary Hospital in Dhaka – An Observational Study. Eur J Med Heal Sci. 2020;2(6):1–22. doi:10.24018/ejmed.2020.2.6.599

30. Ahmed S, Karim MM, Ross AG, et al. A five-day course of ivermectin for the treatment of COVID-19 may reduce the duration of illness. Int J Infect Dis. 2021;103:214–216.

31. Caly L, Druce JD, Catton MG, Jans DA, Wagstaff KM. The FDA-approved drug ivermectin inhibits the replication of SARS-CoV-2 in vitro. Antiviral Res. 2020;178:104787. doi:10.1016/j.antiviral.2020.104787

32. Sharun K, Dhama K, Patel SK, et al. Ivermectin, a new candidate therapeutic against SARS-CoV-2/COVID-19. Ann Clin Microbiol Antimicrob. 2020;19(1):1–5. doi:10.1186/s12941-020-00368-w

33. Chang GA, Figueredo ANT. COVID-19: IVERMECTIN PROPHYLAXIS IN ADULT CONTACTS. Report on Health Personnel and Post-Exposure Prophylaxis. Preprint. Res Gate. 2020;(October). doi:10.13140/RG.2.2.11985.35680/3

34. Wang T, Du Z, Zhu F, et al. Comorbidities and multi-organ injuries in the treatment of COVID-19. Lancet. 2020;395(10228):e52. doi:10.1016/S0140-6736(20)30558-4

35. Renu K, Prasanna PL, Valsala Gopalakrishnan A. Coronaviruses pathogenesis, comorbidities and multi-organ damage – A review. Life Sci. 2020;255:117839. doi:10.1016/j.lfs.2020.117839

